# Body mass index and heart failure risk: a cohort study in 1.5 million individuals and Mendelian randomisation analysis

**DOI:** 10.1101/2020.09.23.20200360

**Authors:** R. Thomas Lumbers, Michail Katsoulis, Albert Henry, Ify Mordi, Chim Lang, Harry Hemingway, Claudia Langenberg, Michael V. Holmes, Naveed Sattar, on behalf of the HERMES Consortium

**Author notes:** **Address for correspondance** Dr R. Thomas Lumbers, Institute of Health Informatics, University College London, 222 Euston Road, London NW1 2DA, United Kingdom. Joint first authors (R. Thomas Lumbers, Michail Katsoulis). Joint last authors (Michael V. Holmes, Naveed Sattar).

## Abstract

**Graphical abstract:** 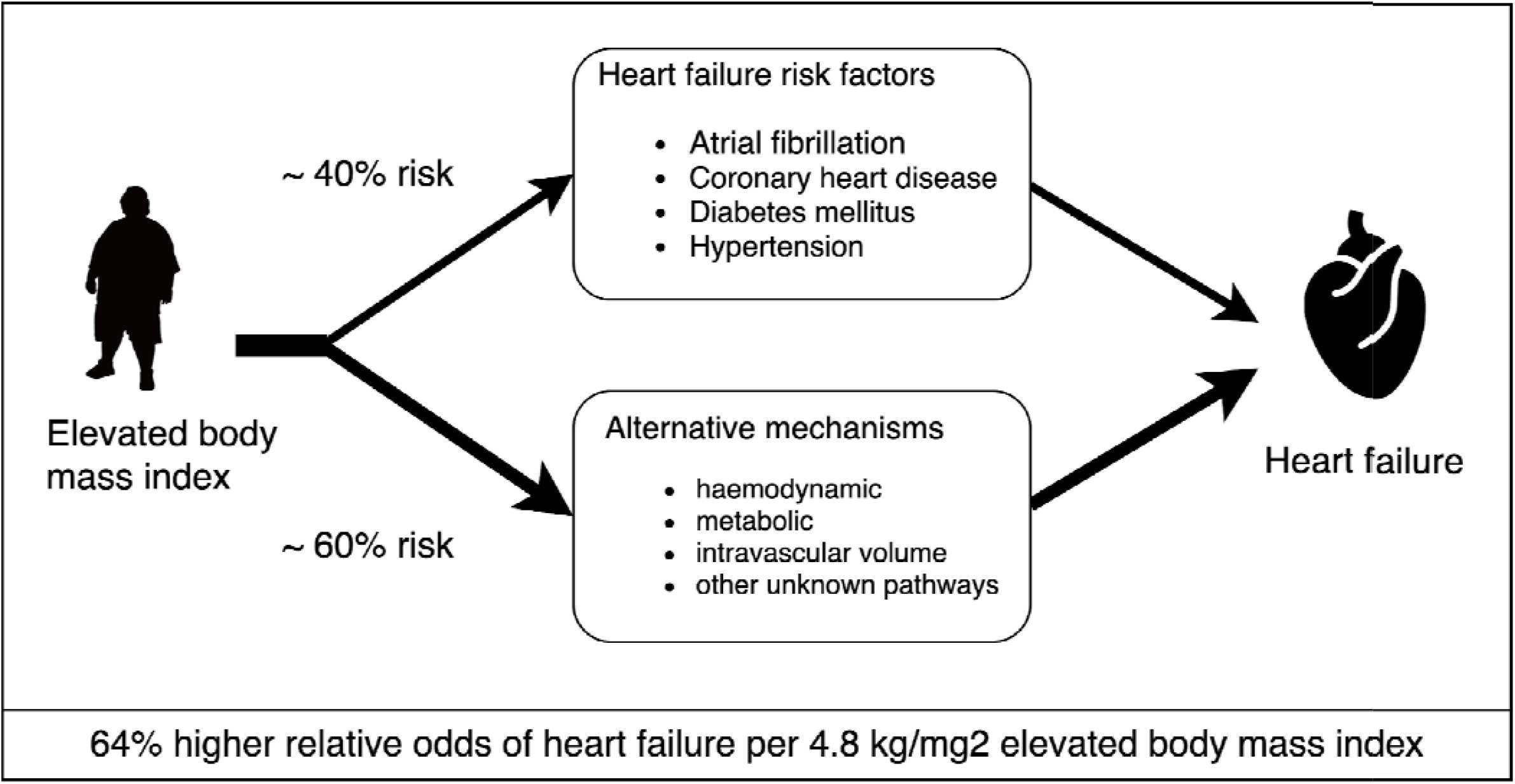

**Aims:** Elevated body mass index (BMI) is a known risk factor for heart failure (HF), however, the underlying mechanisms are incompletely understood. The aim of this study was to investigate the role of common HF risk factors as potential mediators.

**Methods and Results:** Electronic health record data from primary care, hospital admissions and death registrations in England were used to perform an observational analysis. Data for 1.5 million individuals aged ≥18 years, with BMI measurements and free from heart failure at baseline, were included between 1998 and 2016. Cox models were used to estimate the association between BMI and HF with and without adjustment for atrial fibrillation (AF), diabetes mellitus (DM), coronary heart disease (CHD), and hypertension (HTN). Univariable and multivariable two-sample Mendelian randomisation was performed to estimate causal effects.

Among non-underweight individuals, BMI was positively associated with HF with a 1-SD (∼ 4.8kg/m^2^) higher BMI associated with a hazard ratio (HR) of 1.31 (1.30, 1.32). Genetically predicted BMI yielded a causal odds ratio (OR) of 1.64 per 4.8 kg/m^2^ BMI (1.58, 1.70) which attenuated by 41% (to OR of 1.38 (95% CI 1.31-1.45), when simultaneously accounting for AF, DM, CHD and SBP.

**Conclusion:** About 40% of the excess risk of HF due to adiposity is driven by SBP, AF, DM and CHD. These findings highlight the importance of the prevention and treatment of excess adiposity and downstream HF risk factors to prevent HF, even in people in whom the above risk factors are well managed.

**One-sentence summary:** This study of the role of excess adiposity as a risk factor for HF, including an observational analysis of measured BMI 1.5 million individuals and multivariable MR analysis of genetically elevated BMI, provides evidence that adiposity is causally associated with HF, with approximately 40% of the effect being mediated by conventional risk pathways.

## Introduction

Heart failure (HF) affects an estimated 30 million individuals worldwide and is a leading cause of mortality and morbidity.^1^ Understanding the aetiology of HF is essential to inform on therapeutic and lifestyle approaches to minimise the burden of HF-related morbidity and mortality. While HF may share risk factors with CHD, there is emerging evidence that HF also has a distinct aetiopathology, motivating the need for a thorough exploration of potential modifiable risk factors.

The prevalence of overweight and obesity is rising, particularly among lower socioeconomic groups, such that body mass index (BMI) is an important determinant of health inequalities.^2^ Elevated BMI, a surrogate indicator of adiposity, has been associated with a higher risk of incident HF, with the magnitude of association comparatively larger than for myocardial infarction.^3^ Investigating whether this association has a causal basis poses a translational challenge: evidence from randomised controlled trials of weight loss interventions is, as yet, limited to effects on cardiovascular risk factors^4^ and the effects on heart failure risk are unknown.^5^ An improved understanding of the mediating mechanisms by which excess adiposity leads to higher risk of HF may inform therapeutic strategies for prevention in the overweight and obese population, including weight loss interventions.

Evidence from Mendelian randomisation studies suggest that elevated BMI increases risk of HF and diseases that are linked to HF, including, atrial fibrillation (AF)^6^, coronary heart disease (CHD)^7^, diabetes mellitus (DM)^8^ and hypertension (HTN)^9^; for the latter two factors, weight loss trials confirm causal effects^10,11^; however, what is currently unknown is the extent to which the BMI to HF relationship is mediated by these factors. Such knowledge could help inform the targeting of treatments to reduce HF risk in overweight and obese individuals, as a complement to weight loss strategies.

In this study we combine evidence from observational and Mendelian randomisation analysis to explore the extent to which the BMI-HF relationship is mediated by HF risk factors that are the causal consequence of elevated BMI. First, we made use of a large observational sample of individuals to estimate the overall (total) association of BMI on HF as well as the association of BMI on HF independent of the selected risk factors (direct effect); and, second, we estimate equivalent total and direct effects of BMI using univariable and multivariable Mendelian randomisation. Our findings are relevant to HF preventative efforts as they quantify the extent to which the causal effects of BMI on risk of HF might be off set by intervening on downstream sequelae (e.g. HTN, DM) for which effective pharmacological therapies are available. Furthermore, any residual direct effects of BMI not accounted for by the recognised risk factors motivate new avenues of aetiological investigation and highlight the potential for new pharmacological approaches to preventing HF. They would also suggest weight loss may have benefits in prevention of HF even when existing risk factors are well managed.

## METHODS

### Epidemiological association between body mass index and heart failure

#### Data source

The study population was derived from the CALIBER program (CArdiovascular disease research using LInked Bespoke studies and Electronic health Records), a data platform that provides access to longitudinal linked electronic health records in England: primary care records from general practices participating in the Clinical Practice Research Datalink (CPRD); coded discharge data from secondary care from Hospital Event Statistics (HES); and death registrations from the Office of National Statistics (ONS). ^12^ Approval for the study was granted by the Independent Scientific Advisory Committee (ISAC) of the Medicines and Healthcare products Regulatory Agency.

#### Procedures and outcomes

We defined disease status at baseline with respect to HF, AF, CHD, DM on the basis of coded diagnoses recorded in CPRD, HES or ONS (Table S1). Procedures for the development and validation of these disease phenotypes and traits are reported elsewhere (http://caliberresearch.org/).^13^ For baseline BMI, we obtained height, weight, and recorded BMI measurements recorded in CPRD in the year before study entry. In some cases, BMI was recorded, but height and weight measurements were not. Where possible, we calculated BMI as weight/height^2^ (kg/m^2^), or, alternatively used reported BMI measures directly. We excluded individuals with discrepant values for height, weight, and calculated BMI (absolute difference between recorded and calculated BMI, on the same day, > 1kg/m^2^).

#### Study population

Individuals of 18 years or older, between January 1, 1998, and June 30, 2016, with at least 1 year of up-to-standard data in a CPRD practice were identified (N=2,396,229). The following exclusions were made: 64,899 women who were pregnant at baseline; 1080 who had inconsistent data of BMI and weight; 40439 with prevalent HF, 571782 of European ancestry; and 134789 without data on index of multiple deprivation. After the exclusions, 1,583,240 individuals were included in an initial exploratory analysis (Figure S1). We observed a J-shaped association between BMI and HF, with the non-linear component of the association being attributable to underweight individuals (BMI <18.5 kg/m^2^), who contributed only 2.6% of the overall sample. For individuals of normal weight and above (i.e. individuals with BMI ≥ 18.5kg/m^2^), higher BMI was associated with higher risk of HF with the relationship being log-linear; whereas, for underweight individuals (BMI <18.5 kg/m^2^), lower BMI was associated with higher risk (Figure S2). Given that our aim from the observational analysis was to estimate the average effect of BMI across the population distribution without incurring bias, and that the increased risk of HF among those with very low BMI could arise from reverse causation, we excluded individuals who were underweight (BMI < 18.5kg/m^2^).^14^ After exclusions, 1,542,231 individuals were included in the study (Figure S1).

#### Statistical analysis

Proportional hazards models were used to estimate the association between BMI levels and incident heart failure. In our primary analysis, we adjusted for sex and age at enrolment and stratified by general practice. We then performed stepwise analyses adjusting for prevalent AF, CHD, DM, HTN. In sensitivity analysis, we additionally adjusted for potential confounding by social deprivation using the Index of Multiple Deprivation, a score derived from indices such as income, employment, and education. ^15^ We estimated hazard ratios for BMI as both a continuous and categorical variable, using clinically relevant cut-off points: normal weight (BMI 18.5 - 24.9kg/m^2^), overweight (BMI 25-29.9 kg/m^2^), moderately obese (BMI 30 - 34.9 kg/m^2^), severely obese (BMI 35 - 39.9 kg/m^2^), very severely obese (BMI ≥ 40 kg/m^2^).^16^ As described above, we excluded underweight individuals (BMI < 18.5kg/m^2^), because we wanted to avoid bias due to reverse causation. To enable comparison with genetic association data, hazard ratios were calculated per 4.807 kg/m^2^, corresponding to the standard deviation of BMI within the UK Biobank samples.^17^ The analyses were conducted using StataMP 16 (StataCorp, College Station, TX, USA).

### Characterising the relationship between body mass index and heart failure, and exploring potential mediation, by Mendelian randomisation

#### Data sources and selection of instrumental variables

Genetic association estimates for the exposure (BMI), outcome (HF), and potential mediators (AF, CHD, DM, and systolic blood pressure (SBP)) were obtained from published genome-wide association studies (GWAS), (Table S2).

We developed a linkage disequilibrium reference dataset based on 10,000 unrelated individuals of European ancestry, selected at random from UK Biobank participants, with genotypes imputed to the Haplotype Reference Consortium.^18^ Based on genetic quality control (QC) metrics provided by UK Biobank, participants were excluded based on sex mismatch between submitted and inferred sex, having more than 10 putative third-degree relatives, excessive heterozygosity and missing rates, withdrawal from study, and missing QC metrics. Variants with imputation info score <0.3, minor allele frequency outside of 0.01 - 1 range, and missing call rate ≥ 5% were excluded. This results in a set of 408,480 individuals from which a random sampling without replacement was used to extract unrelated individuals for creating the LD reference panel.

To identify independent genetic instrument variables for the exposure, and potential mediators, variants associated with each trait at conventional levels of GWAS-significance (*P* < 5 × 10^−8^) were pruned for linkage disequilibrium, using *r*2 < 0.01.

#### Univariable Mendelian randomisation analysis

The primary Mendelian randomisation analysis was performed using two-sample, inverse-variance weighted meta-analysis, under a random effects model. The published GWAS summary estimates for BMI were provided for a rank-based inverse normal transformation of the phenotype, therefore, the beta coefficients correspond, approximately, to effect sizes per standard deviation increase of genetically estimated BMI (4.807 kg/m^2^).^17^ All Mendelian randomisation analyses were conducted using the MendelianRandomisation package^19^ in R (R Foundation).

We performed a series of sensitivity analyses to investigate whether there was evidence of directional (i.e. unbalanced) horizontal pleiotropy and/ or invalid instruments. First, we derived parameter estimates using the weighted median approach; this method can provide valid estimates where the weight of up to half of the instruments is invalid (i.e. does not meet the assumptions required for valid Mendelian randomisation).^20^ Second, we used the MR Pleiotropy RESidual Sum and Outlier (MR-PRESSO) method to identify and exclude potential outlier instrument variables^21^. Finally, we applied the MR-Egger method.^22^

#### Multivariable Mendelian randomisation for mediation

To estimate the direct effects of BMI on HF after taking into account the mediating effects of SBP, AF, CHD and DM, we employed two complementary approaches. In our main analysis, we first selected instruments for each exposure trait as described above (i.e. SNPs associated with the trait at GWAS significance P < 5×10-8 with a between-SNP correlation of r2 < 0.01). Where genetic variant instruments for two or more traits were in higher linkage disequilibrium than that which we used to prune SNPs entering our instruments (i.e. when *r*^2^ > 0.01), we selected the variant with the smallest association *P* value for BMI (given that BMI was our primary exposure). For each BMI and risk factor combination, we created a joint instrument set using the genetic association estimates with each trait under analysis. This instrument set was used in a multivariable Mendelian randomization analysis with inverse-variance weighted ^23^ and MR-Egger regression^24^ to estimate the direct causal effect of BMI accounting for other traits in the model. Further, to test the extent to which these estimates were affected by prioritisation of instrument to BMI association, we reran the analyses excluding SNPs at random (rather than prioritising on the BMI association) where SNPs were correlated (LD *r*^2^ > 0.01) in joint instruments. Using these approaches, we tested the direct causal effect of BMI with single covariate adjustments on: 1) SBP, 2) AF, 3) CHD, and 4) DM and multi-covariate adjustments on 5) AF-CHD-DM-SBP. As a sensitivity analysis, we repeated the multivariable Mendelian randomisation analysis using an alternative method whereby we performed univariable MR with stepwise conditioning of the outcome dataset for each risk factor (and all risk factors in combination), using mtCOJO.^25^

#### Participant overlap in two-sample Mendelian randomisation

There is potential participant overlap between the UK Biobank + GIANT GWAS of BMI (total sample 694,649, of which 484,680 were from UKB) and the HERMES GWAS of HF (6,504 cases, 47,309 controls), equating to a potential case overlap of only 6,504/694,649=0.9% and therefore a negligible risk of bias leading to type 1 error.^26^

## RESULTS

### Body mass index and risk of incident heart failure

Our observational analysis consisted of 1,542,231 individuals following the exclusion of 41,009 subjects with BMI <18.5 kg/m^2^ (see Methods for further details). The mean (SD) age was 51.1 (18.3) years old and 60% of participants were female. The mean (SD) BMI at enrolment was 27.4 (5.6) kg/m^2^ which was similar to that reported for the UK Biobank [27.4 (4.8)]. Descriptive characteristics for the study population are given in **Table 1**. Of the individuals included in the study, 5.2% had atrial fibrillation, 9.7% CHD, 6.1% DM, 26.6% HTN, at baseline. The median follow-up time was 6.1 (IQR=2.7-10.0) years and during this period, 65,918 participants (4.1%) were diagnosed with heart failure.

**Table 1:** Characteristics of study population at baseline, according to body mass index category.

|  | **Normal weight**<br>**(BMI 18.5-24.9**<br>**kg/m<sup>2</sup>)** | **Overweight**<br>**(BMI 25-29.9**<br>**kg/m<sup>2</sup>)** | **Obese I**<br>**(BMI 30-34.9**<br>**kg/m<sup>2</sup>)** | **Obese II**<br>**(BMI 35-39.9**<br>**kg/m<sup>2</sup>)** | **Obese III**<br>**(BMI ≥40**<br>**kg/m<sup>2</sup>)** |
| --- | --- | --- | --- | --- | --- |
| **N** | 586,370 | 547,268 | 265,202 | 95,297 | 48,094 |
| **Follow-up (years)** | 6.4 ± 4.5 | 6.8 ± 4.5 | 6.8 ± 4.5 | 6.7 ± 4.6 | 6.5 ± 4.5 |
| **Age (years)** | 48.7 ± 20.1 | 53.8 ± 17.3 | 52.4 ± 16.3 | 49.2 ± 15.7 | 46.3 ± 14.7 |
| **Female sex: N (%)** | 399,992 (68) | 282,549 (52) | 145,343 (55) | 61,509 (65) | 35,220 (73) |
| **Coronary heart disease:**<br>**N (%)** | 44,223 (8) | 60,027 (11) | 30,136 (11) | 9,775 (10) | 4,586 (10) |
| **Atrial fibrillation: N (%)** | 26,041 (4) | 31,295 (6) | 15,197 (6) | 4,752 (5) | 2,270 (5) |
| **Diabetes mellitus: N (%)** | 19,212 (3) | 33,387 (6) | 23,632 (9) | 10,523 (11) | 6,559 (14) |
| **Hypertension: N (%)** | 121,591 (21) | 161,624 (30) | 83,234 (31) | 29,432 (31) | 14,376 (30) |

An approximately log-linear association of measured BMI with incident HF was observed for individuals with normal or elevated BMI (**Figure 1**). We estimated proportional hazards for a linear model for BMI and found an increase of 4.807 kg/m^2^ (one SD in UKB) was associated with higher risk of HF (HR 1.31 [95% CI 1.30, 1.32]), (**Figure 2**). To estimate the causal effect of BMI on HF, we performed univariable two-sample Mendelian randomisation using the inverse variance weighted method. This estimated the effect per one SD higher BMI, including those effects mediated through other traits, such as HF risk factors (i.e. the ‘total effect’). The findings from MR supported a causal role of BMI on risk of HF, with an OR of 1.64 (95% CI 1.58, 1.70) (**Figure 3, Table S3**).

**Figure 1.**
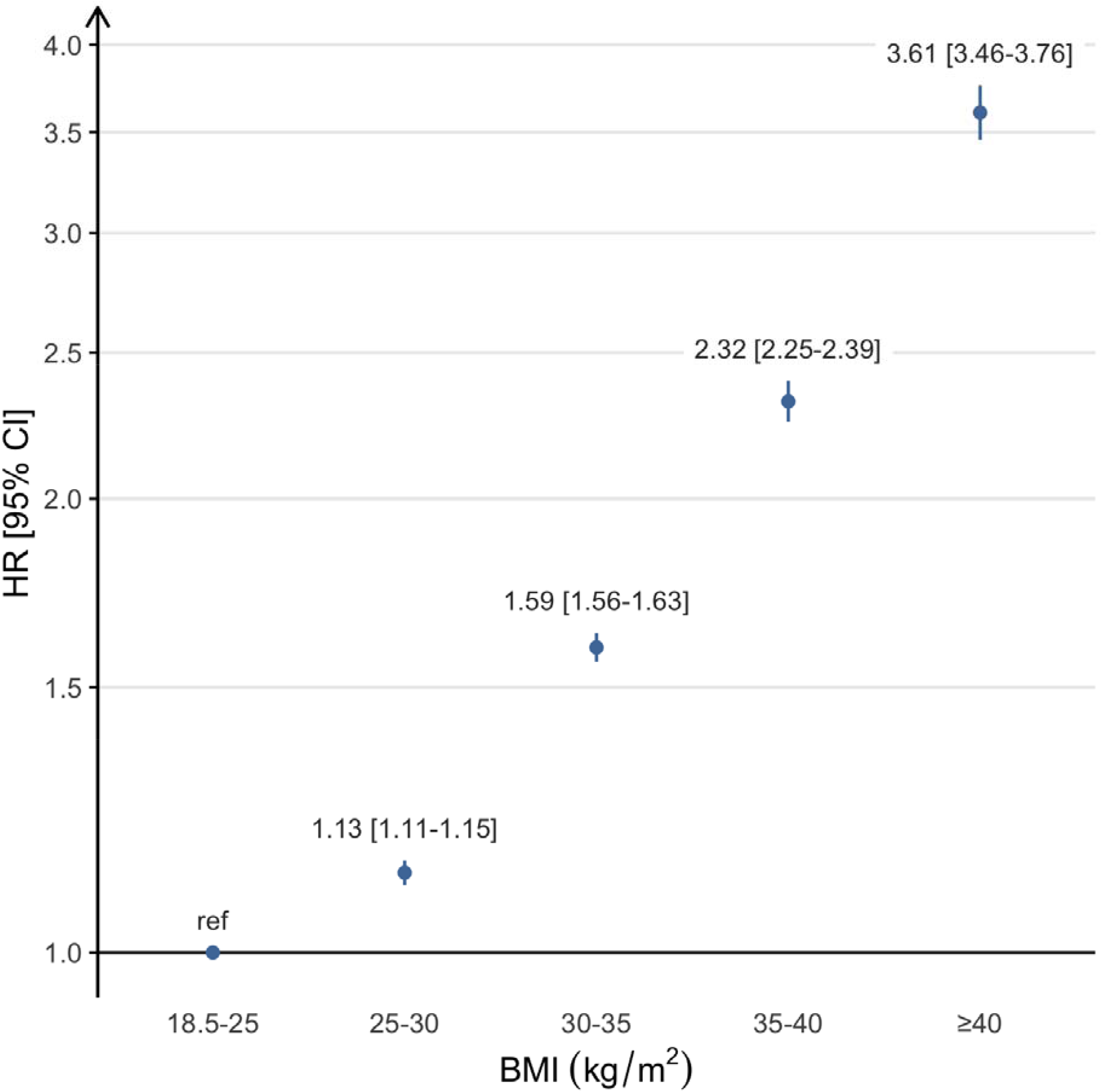
Association of body mass index with incident heart failure according to WHO obesity categories. Estimated hazard ratios for body mass index (BMI) on incident heart failure, adjusted for age and sex according to the World Health Organisation obesity classification. Hazard ratios are given per 4.807 kg/m^2^ increase in BMI, equating to the standard deviation of BMI in UK Biobank.

**Figure 2.**
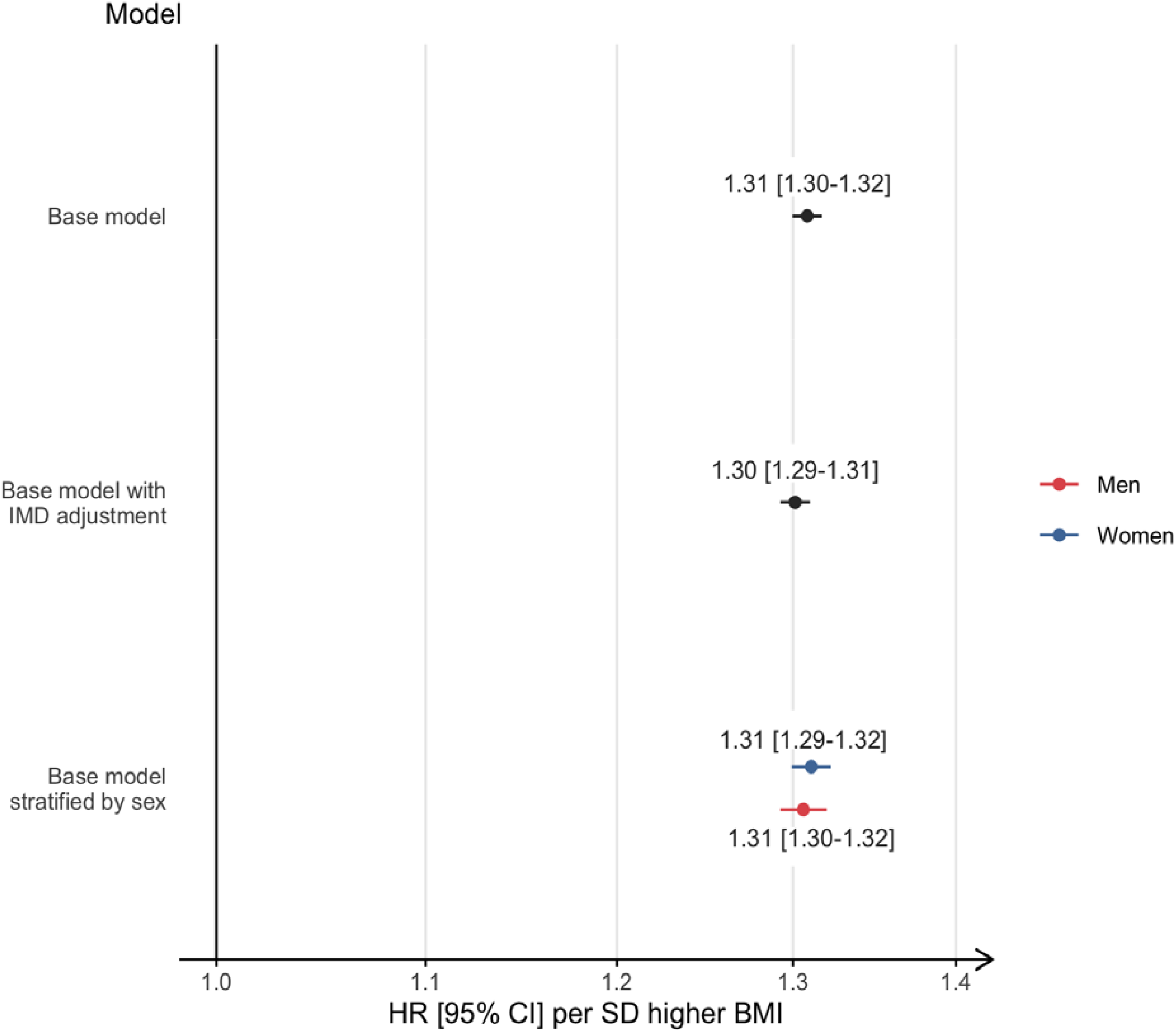
Association of measured body mass index, as a continuous exposure, with incident heart failure. Base models were adjusted for age and sex and additional risk factors. Points and error bars represent adjusted hazard ratios and 95% confidence interval for the effects of body mass index (BMI) on incident heart failure. Hazard ratios are given per 4.807 kg/m^2^ increase in BMI, corresponding to the approximate standard deviation of the sample used for genetic inference analyses. IMD, Index of Multiple Deprivation.

**Figure 3.**
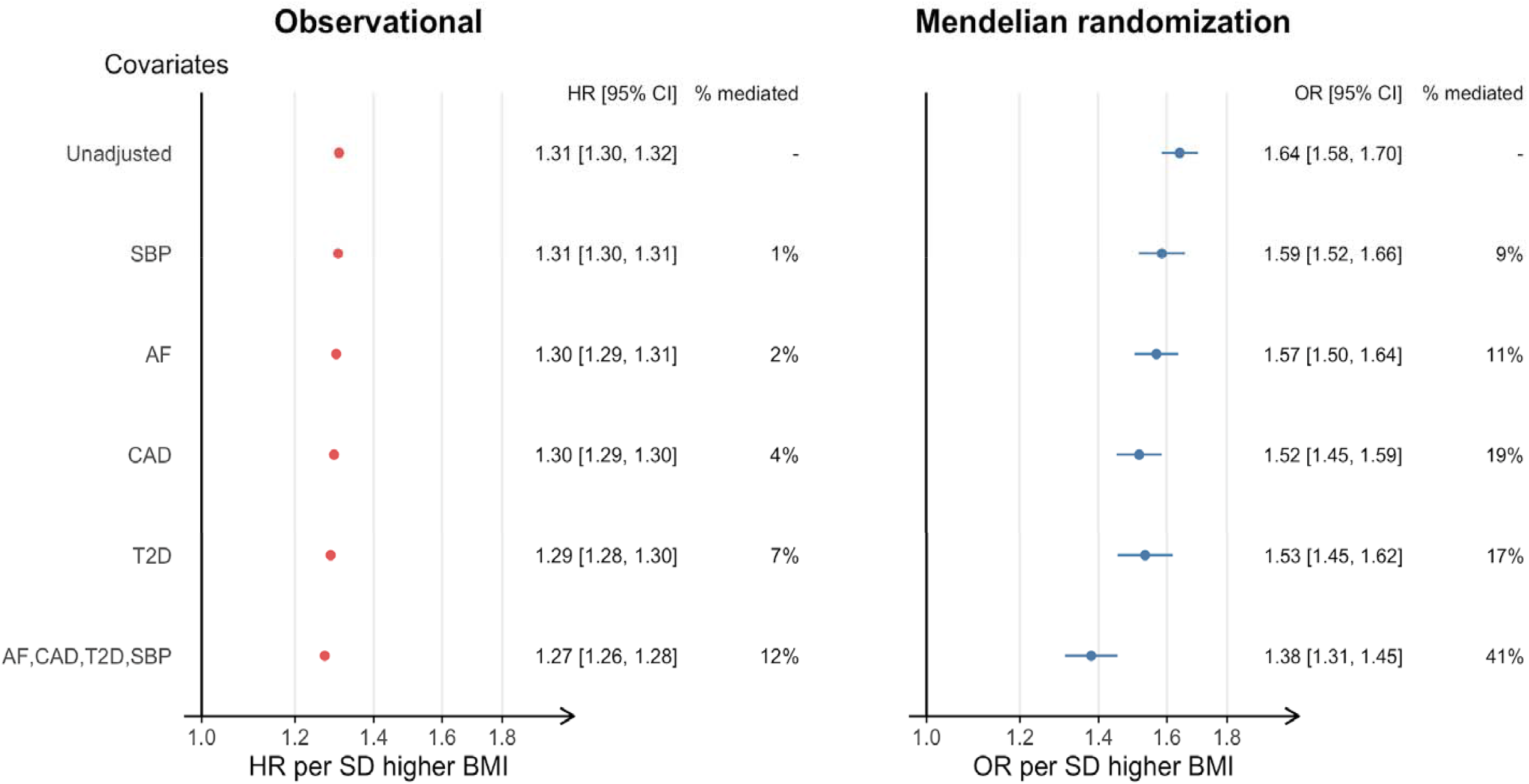
Observational and genetic associations of body mass index with heart failure, with adjustment for sex, age, and risk factors. Left panel: Adjusted hazard ratios for the effects of body mass index (BMI) on incident heart failure, adjusted for age and sex (base model), and additional adjustment for atrial fibrillation (AF), coronary heart disease (CHD), diabetes mellitus (DM), hypertension (HTN). Right panel: odds ratios for the effects of body mass index (BMI) on heart failure, adjusted for age, sex, AF, CHD, DM, systolic blood pressure (SBP), estimated by multivariable Mendelian randomisation. Effect estimates are given per 4.807 kg/m^2^ increase in BMI, equating to the standard deviation of BMI in UK Biobank.

### Mediation of effects of body mass index by heart failure risk factors

Next, we estimated the effects of BMI on HF taking into account potential mediation, using covariate adjustment and multivariable Mendelian randomisation, for four important risk factors for HF [AF, CHD, DM and HTN (SBP in mendelian randomisation)] according to their reported prevalence and population attributable risk^27^. These analyses allowed us to estimate the direct effects of BMI on HF, i.e. independent of the risk factors of interest.

Using observational data, we repeated the proportional hazards estimation with adjustment analysis for each risk factor in turn, and for all factors combined (**Figure 3, left panel**). The effect of adjustment of all baseline factors combined was modest [HRadj 1.27 per 4.807 kg/m^2^ increase (95% CI 1.26, 1.28)], in other words the indirect effect through these factors was only 13% of the overall effect of BMI on risk of HF.

We then performed two-sample multivariable Mendelian randomisation to estimate the direct effects of BMI, accounting for potential mediators, based on genetic predisposition (hereafter referred to as adjusted effects). We found that the OR estimate attenuated upon adjustment for each of the risk factors of interest [1.64 unadjusted (CI 1.58, 1.70) to 1.38 (CI 1.31,1.45) fully adjusted], suggesting that 41% of the total effect of BMI on HF is mediated by these factors (**Figure 3, right panel**).

### Sensitivity analyses

We performed sensitivity analyses to assess the effects of social deprivation as a potential confounder of the observational association between BMI and HF. The results were robust to adjustment with minimal attenuation of effects after adjustment (**Figures 1 and Figure S3**). To mitigate the possibility that some subjects may have pre-clinical or undiagnosed HF at the time of enrolment, we excluded individuals in whom a diagnosis of HF was made within two years of follow up and found no material change to the estimates (**Figure S3, panel C**).

We performed a series of sensitivity analyses to evaluate the robustness of our Mendelian randomisation. The univariable and multivariable MR-Egger analysis did not indicate directional pleiotropy and revealed consistent estimates, but with lower precision, as expected. Similar estimates to the inverse-variance weighted model we obtained from the weighted median estimator (**Table S3**). The MR-PRESSO method identified up to four outliers for different combinations of covariates in the model, however outlier-correction did not materially change the parameter estimates [OR for BMI 1.65 (95% CI: 1.58-1.73)] (**Table S4**). We used an alternative method, mtCOJO, to multivariable Mendelian randomisation to quantify the potential mediation using and the findings were comparable results (**Table S5**).

## DISCUSSION

### Principle findings

We conducted an observational and MR analysis to evaluate the role of adiposity in the development of HF and to investigate the role of established HF risk factors, AF, CHD, DM and SBP in mediating these effects. Consistent with previous studies, the results from our observational analysis demonstrate a continuum of risk associated with excess adiposity with an approximately log-linear association between higher BMI and HF risk among overweight or obese individuals. Using genetic association data and MR analysis we demonstrate that this relationship is likely to be causal and estimate that ∼40% of the BMI effect on HF is mediated through these established HF risk factors. In contrast to other cardiovascular diseases, such as stroke and coronary artery disease, for which the majority of the BMI risk effects are mediated through conventional risk factors^28^, our findings suggest an important role for alternative pathways to HF. They highlight the importance of maintaining a healthy body weight to prevent HF, even in people in whom established HF risk factors are well managed. The findings also suggest some merit in considering intensive weight loss as an intervention in very obese patients with symptomatic HF despite appropriate standard of care medical therapy.

### Comparison with other studies

Our study of the association of measured BMI and the incidence of HF in ∼ 1.5 million individuals demonstrated a positive dose-response relationship, with higher hazards of 31% per standard deviation increment (equivalent to 4.807 kg/m^2^). These findings are consistent with estimates from a meta-analysis of 647, 388 participants from 23 prospective studies which estimates a relative risk of 1.41 per 5 kg/m^2^ higher BMI^29^. Our estimate of the causal effect of BMI on HF, based on MR analysis, was of comparatively greater magnitude (OR = 1.64), under the assumption that hazard ratios and odds ratios approximating to risk ratios given the outcome is rare^30^; these findings are in line with previous MR studies.^31^ This larger effect may be explained, in part, by the lifelong and cumulative exposure to genetically elevated BMI; interestingly, a previous study reported an association between of measured BMI in young adults and incident HF that was of greater magnitude relative to studies of older populations.^3^ Furthermore, BMI in older people is also more prone to be influenced by reverse causality whereby underlying disease, whether diagnosed or not, can lead people to lose weight or slow weight gain. Our observation and MR analyses, through triangulation, provide strong evidence that excess adiposity is causally related to the development of HF. We are not aware of any previous MR studies evaluating the role of common HF risk factors, AF, CHD, DM and SBP, as mediators of the effects of excess adiposity on HF.

### Clinical implications

Our results confirm and extend the results of previous studies and demonstrate that excess adiposity is an important causal factor for HF. Interestingly, the population attributable risk for HF was nearly equivalent to that for hypertension, even at times of lower prevalence rates of overweight and obesity.^32^ Hypertension and CHD have been considered the principle risk factors for HF and the main focus of therapeutic interventions. A recent meta-analysis of the effects of anti-glycaemic therapies on HF risk in patients with diabetes found evidence that weight change was an important effect modifier, consistent with a causal role for excess adiposity.^33^ Taken together, these findings support a causal role for excess adiposity and suggest that, in line with current treatment guidelines for weight loss interventions to delay or prevent HF, weight loss reduction therapies are likely to be an important component of strategies to address the growing epidemic of heart failure.^5^

We find that SBP, DM, CHD, and AF, established HF risk factors for which effective treatments exist, play an important role as mediator of the BMI to HF relationship suggesting that effective treatment may mitigate risk. Conversely, our results highlight the importance of additional disease mechanisms that may, potentially, account for the residual causal effects of BMI on HF (even when taking into account the risk factors we studied). One such mechanism may be the haemodynamic dysregulation and increased plasma volume status that is observed in obesity, which is reversible with weight loss, and which may contribute to the development of HF.^34^ Indeed, the benefit of SGLT2i on the prevention of HF in diabetes populations, with high prevalence of overweight and obesity, has been considered to occur, at least in part, via improvements in plasma volume status and other mechanisms linked to altered nutrient signalling.^35,36^ Other proposed mechanisms include increased cardiac work; sleep disordered breathing and increased pulmonary arterial pressures; and the adverse effects of obesity-driven systemic inflammation.^37^

### Strengths and limitations

The key strengths of this study include the large study sample, both for the observational and MR analyses, which allowed us to generate precise estimates of the effects of BMI on HF and provided sufficient power for adjusted, multivariable and subgroup analysis. A further strength is that we limited the study population to individuals of European ancestry thus reducing any potential bias due to population stratification. An important limitation in our estimates of the direct and indirect effects of measured BMI on HF in the observational analysis is that we only considered HF risk factors present at baseline, and not through follow-up, which may have led to an underestimation of the mediating effects. In addition, we did not subclassify HF cases by left ventricular ejection fraction or predominant aetiology and it is possible BMI effects are differential for particular disease subtypes.

The MR analysis approach employed here mitigates for the effects biases due to confounding and reverse causation that impact all studies using measured BMI to evaluate the relationship between excess adiposity and downstream clinical outcomes.^38^ Additionally, the use of multivariable MR enabled us to quantify the mediating effects of BMI-associated HF risk factors for the BMI to HF relationship and to demonstrate the extent of the ‘residual’ direct effects of BMI, which represent a potential opportunity for the discovery of new aetiological pathways and therapeutic targets.^39,40^

### Future directions

This study provides important information by estimating the risk effects of excess adiposity and exploring the mediating role of conventional HF risk factors, AF, CHD, DM and SBP. The majority of the HF risk associated with elevated BMI was not associated with these risk factors and there is a need for further research to explore these alternative mechanisms. In particular, the effects on myocardial structure and function warrant further study. Finally, there is some evidence to suggest that excess adiposity is a determinant of adverse outcomes in people with prevalent HF ^41,42^ and there is a need for clinical outcomes trials to evaluate the safety and efficacy of specific weight loss interventions.

## Conclusion

This comprehensive appraisal of the role of excess adiposity as a risk factor for HF, including an observational analysis of 1.5 million individuals and multivariable MR analysis, provides evidence that higher BMI is causally related to higher risk of HF. The MR analysis suggests that ∼ 40% of the effect of BMI on HF is mediated by conventional risk pathways. Our work would suggest that weight loss may lessen HF risk over and above optimal treatment of these factors and point to the need to test the effectiveness of specific interventions in randomised controlled trials both for the prevention and treatment of HF.

## Data Availability

Data is not available

